# The design of an orthodontic patient information leaflet: a cross-sectional, mixed methods study involving surveys, a knowledge test and eye-tracking

**DOI:** 10.64898/2026.10.25.26366089

**Authors:** Damian JJ Farnell, Aik Jin Kho, Jennifer Galloway

## Abstract

**Background:** Patient Information Leaflets (PILs) are widely used to support verbal communication during orthodontic consent. For a PIL to be effective, it must present information clearly and support patient understanding and recall. A commonly used orthodontic PIL in the UK is the British Orthodontic Society’s (BOS) PIL “Orthodontic Treatment: What are the risks?”.

**Aim and objectives:** To evaluate this orthodontic PIL, compared to a simpler Designed to Smile PIL, with regards to: 1) Design clarity; 2) Free text comments on the design and content; 3) Knowledge after reading the PIL; 4) Time spent looking at various aspects of the PIL.

**Methods:** Students of non-medicine or dentistry programmes viewed the orthodontic PIL electronically with simultaneous eye-tracking. Immediately afterwards, they completed a survey capturing their clarity of design rating, feedback on the design and content, and knowledge of the PIL content. This was repeated for a simpler Designed to Smile PIL.

**Results:** 36.8% rated the orthodontic PIL as “very clear” compared to 66% for the Designed to Smile PIL. Suggested improvements included breaking-up large blocks of text, using more images, and including descriptions of patient experience. 48.9% (SD = 9.0) of questions were answered correctly regarding content in the orthodontic PIL; significantly lower than the simpler Designed to Smile PIL (80%; SD = 11.3%) (*P* < 0.001). Eye-tracking indicated that participants spent more time looking at images in the orthodontic PIL (median = 22.2% of time) compared to the Designed to Smile PIL (median = 8.2%) (*P* < 0.001).

**Conclusions:**

- The Designed to Smile PIL was deemed clearer than the orthodontic PIL.
- Suggestions for improvement included using more images and breaking-up large sections of text.
- Knowledge was relatively poor after reading the orthodontic PIL.
- Participants spent longer looking at the images in the orthodontic PIL compared to the Designed to Smile PIL.

## Introduction

### Background

Severe malocclusion has been shown to have a negative impact on oral health-related quality of life (Chiba *et al*., 2022). Patients undergoing orthodontic treatment are often motivated by a desire for an improvement in appearance alongside function (Burden *et al*., 1995; Tulloch *et al*., 1984). This may in turn improve self-esteem and help with social interactions, ultimately having a positive psychological impact on the patient (Grewal *et al*., 2019; Mandava *et al*., 2021).

However, orthodontic treatment has associated risks. Common risks include demineralisation (Benkaddour *et al*., 2014), gingivitis (Boke *et al*., 2014), root resorption (Sameshima *et al*., 2021), and pain and discomfort (Long *et al*., 2016; Ngan *et al*., 1989). It is therefore crucial that information about risks is given to orthodontic patients prior to starting treatment to ensure adequate consent.

Effective communication during the consent process is important to ensure consent is valid. Patient Information Leaflets (PILs) can help during this process (Brown *et al*., 2004) and may also improve patient satisfaction with treatment (Ley, 1982). The use of PILs alongside verbal information has been shown to increase patients’ recall of advice (Gauld, 1981), which is essential for adequate consent. Research has also shown that patients are supportive of receiving written information, with 75% of patients reporting that they wanted written information and 80% reporting reading the leaflets given (Weinman, 1990). Written information has the additional benefit of being available for patients to reread the information at home (Bishop *et al*., 1997).

However, to be effective, PILs need to be suitably designed to facilitate knowledge transfer. Evidence exists relating to effective design of PILs including: giving consideration to font size, using short sentences, using short blocks of text in paragraphs, using simple language, ensuring suitable readability scores, being accessible to all patients, emphasising key points, and using colour (Adepu *et al*., 2012; Dowse *et al*., 2011; Hemphill, 1996; Jansen *et al*., 2008; Karreman *et al*., 2007; Kitching, 1990; Pires *et al*., 2015; Pothier *et al*., 2008).

In addition to these aspects of PIL design, another important technique is the use of images to convey information. Supplementing written text with illustrations has been shown to improve patient information recall (Brotherstone *et al*., 2006; Mepplink *et al*., 2015). The addition of pictures has also facilitated understanding in patients with poorer literacy skills (Dowse *et al*., 2001; Dowse *et al*., 2011; Houts *et al*., 2006) and cartoon illustrations have been shown to improve the readability and compliance of patients (Delp *et al*., 1996). However, the addition of symbols in PILs for participants with learning disabilities may not improve understanding (Poncelas *et al*., 2007) and may even cause confusion (Strydom *et al*., 2001).

The number of studies that have considered the design of orthodontic PILs is relatively small (Harwood *et al*., 2004; Parker, 1996; Patel *et al*., 2008; Seehra *et al*., 2016; Thickett *et al*., 2006). These sources consider similar topics to those presented above for PIL design more generally, including readability (Harwood *et al*., 2004), quality assessment (Seehra *et al*., 2016), and information retention (Patel *et al*., 2008; Thickett *et al*., 2006).

More recently, research has focused on digital resources such as YouTube videos (Liew *et al*., 2025), TikTok videos (Badri *et al*., 2025), animated videos (Shqaidef *et al*., 2021), and audio visuals (Utari *et al*., 2025). Online information was found to be valuable (Wade *et al*., 2025), although there was a danger of “information overload.” AI tools such as Large Language Models (Sokmen *et al*., 2025) and AI-based chatbots (Santonocito *et al*., 2025) are now also used for orthodontic information creation and / or dissemination. However, in the UK, the BOS PILs are currently the most frequently used adjunct to verbal information by orthodontic professionals, alongside practice specific written materials (Williams *et al*, in press).

### Rationale for the Study

As BOS PILs are routinely used throughout the UK to support informed consent, it is important to establish whether they continue to communicate information effectively. Systematic evaluation of their design allows shortcomings to be identified and guides the refinement of materials to better meet patient needs.

### Aim

To extend the body of literature, the overall aim of this study was to evaluate the design of the commonly used BOS PIL “Orthodontic treatment: What are the risks?” and compare this to a simpler PIL, “Healthy Smiles for children age 3 and over”, distributed by Designed to Smile, a dental public health campaign in Wales.

*Primary objective on which the sample size calculation was based:* To evaluate participants’ ratings on the clarity of the design of the PILs.

*Secondary objectives:*

- To evaluate participant’s free text comments on the design of the PILs.
- To test the participants’ knowledge of orthodontic risks after reading the PILs.
- To assess the time participants spent looking at various aspects of the PILs using eye tracking.

## Materials and Methods

### CROSS statement

This report has been prepared in accordance with CROSS criteria for reporting survey studies (Sharma *et al*., 2021).

### Ethical approval

A favourable ethical opinion was granted for this study in April 2022 from the Dental School Research Ethics Committee at Cardiff University (project number: 2204). All participants were given a participant information sheet to keep, which described the aims, remit, and ethical issues involved with this study. Written informed consent to participate in this study was subsequently obtained.

### Study Design

This study used an observational, cross-sectional design including surveys and eye-tracking.

### Participants and Recruitment

A convenience sample of undergraduate male and female students, aged 18 to 21, who were not enrolled on a medical or dental degree were recruited to participate over a three-month period in 2022 as a proxy for a patient population. Participants were recruited via institutional email lists.

### Sample Sizes

A formal sample size calculation was not possible as there was no prior information on which to base this calculation. A statistician (one of the authors) recommended the use of 60 participants following a broad “rule of thumb” recommendation in the literature (Wilson Van Voorhis and Morgen, 2007). However, only 53 people responded to the invitation to participate, which was slightly lower than this *ad hoc* sample size estimate (88.3%). A pilot study was not carried out for these initial investigations.

### Survey development

Two surveys, one survey per PIL, were used (Appendix C) to gather the following information:

- Demographic data, including potential confounding factors:

`o` Orthodontic PIL: Course of study, year of study, sex, visual impairments, previous orthodontic treatment, previously seen the PIL.
`o` Designed to Smile PIL: Course of study, year of study, sex, visual impairments, parent or guardian to any children, previously seen the PIL.
- Participant’s opinions on the clarity of the PIL (5-point Likert scale).
- Open, free text questions related to the PIL design (best features, areas for improvement, missing information).
- A short multiple-choice quiz to assess knowledge after reading the PIL.

Face and content validity of the surveys was assessed and refined through discussion within research group and feedback from members of the multidisciplinary ethics committee. For questions related to PIL design, participants rated the PIL clarity in one question and the further questions were open questions. Where testing knowledge, there are distinct right and wrong answers. In both cases, an underlying trait is not being measured indirectly and so *post-hoc* measures of internal consistency or reliability were not required.

### Data Collection

Data collection was carried out in a private, quiet environment. All data were gathered anonymously, with no identifiable information collected. Data was collected, stored, and analysed in-line with GPDR and Cardiff University data handling guidelines. Participants completed the study for the orthodontic PIL first, and immediately after, repeated the study for the Designed to Smile PIL (Figure 1).

**Figure 1:**
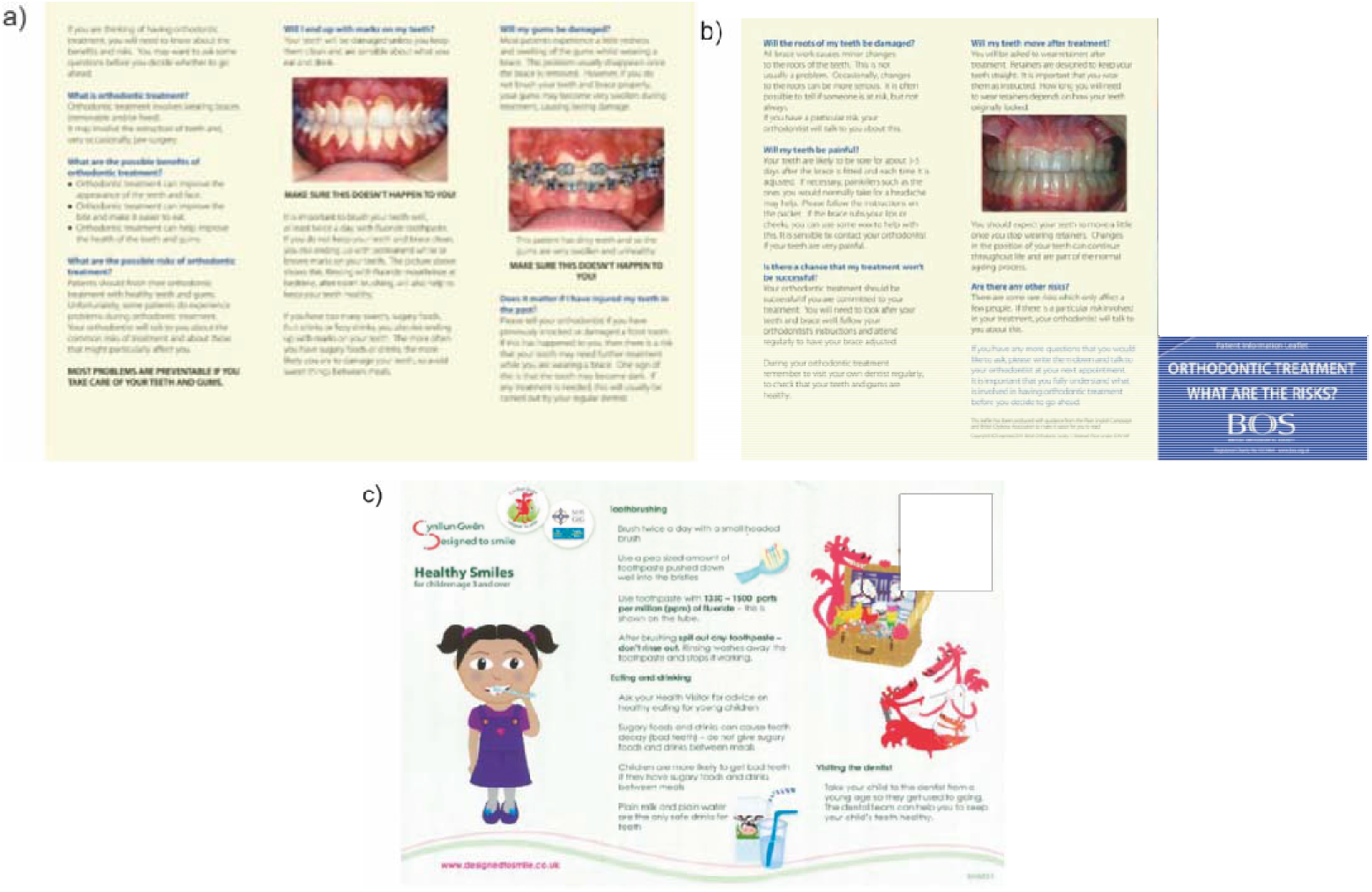
The Patient Information Leaflets (PILs) used in the study (a) British Orthodontic Society’s “Orthodontic treatment: What are the risks?” page 1, (b) British Orthodontic Society’s “Orthodontic treatment: What are the risks?” page 2, and (c) the “Design to Smile” patient information PIL.

The researcher was provided with training prior to undertaking data collection, including set-up, calibration and use of the eye-tracker. Participants were shown the orthodontic PIL on a study specific laptop. To minimise eye-tracking data overwhelm, the first page of the PIL was shown for 75 seconds and then the second page for another 75 seconds. Whilst the participants were reading the PIL, eye movements were monitored using the Tobii© Pro Nano (Figure 2). Further details on the eye-tracking process, including calibration, are available in Appendix A.

**Figure 2:**
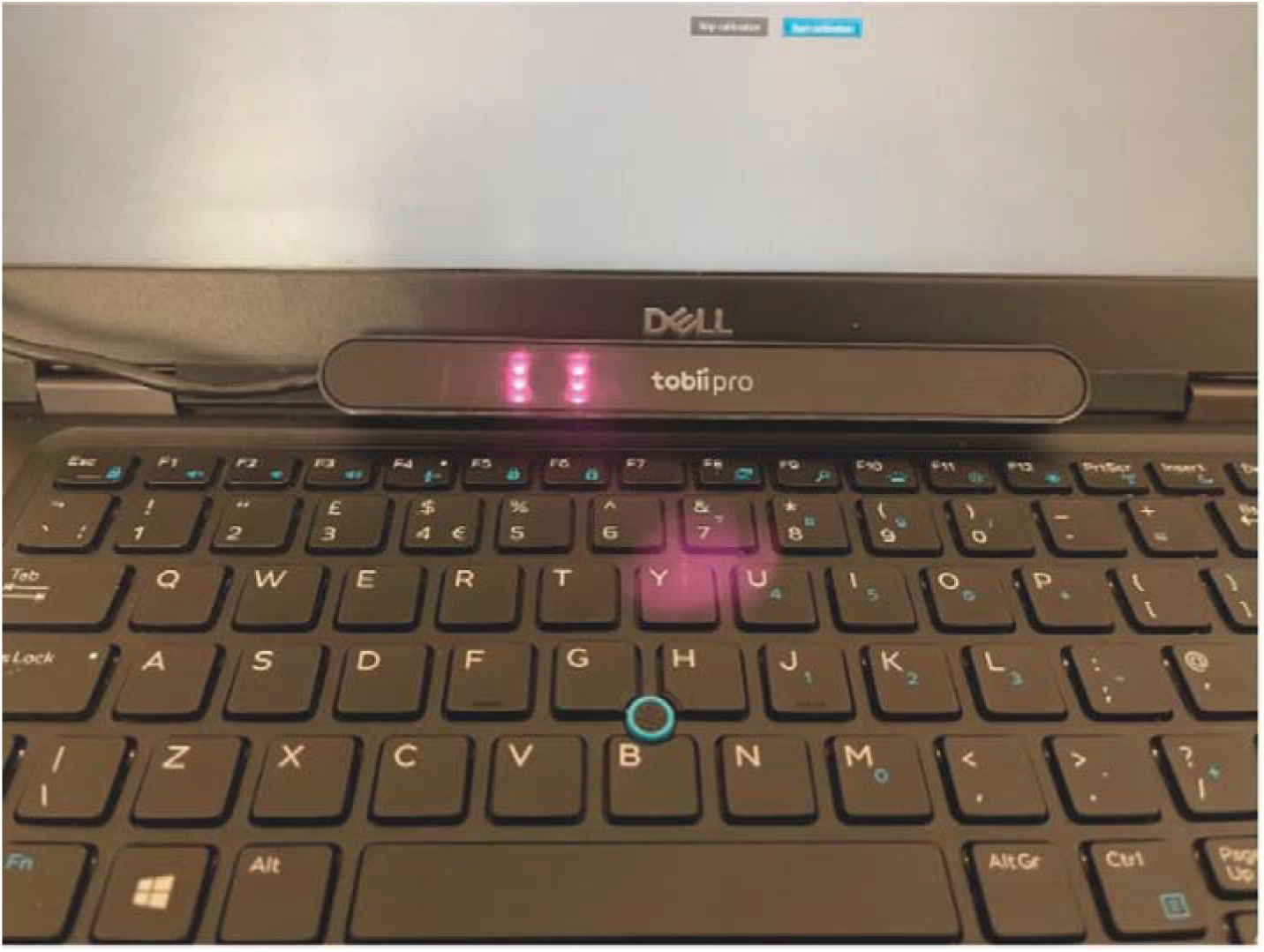
The Tobii© Pro Nano is mounted beneath the screen of a laptop in order to capture eye movements.

Immediately after reading the orthodontic PIL, participants were asked to complete the survey relevant to the PIL (Appendix C). The process was subsequently repeated for the Designed to Smile PIL.

### Data Analysis

#### Eye-tracking data management

“Gaze plots” and “heat maps” of the eye-tracking were generated using the Tobii Pro Lab software. Data from Tobii Pro Lab was subsequently exported to Microsoft Excel. A graphical user interface shown in Figure 3 was created in Visual Basic by one of the authors to help interpret and visualise this data. This interface allowed the user to determine the total number of fixation points and the total time each participant spent looking at specific regions of interest in the PILs. Further information relating to how the user interface was used is presented in the Appendix B.

**Figure 3:**
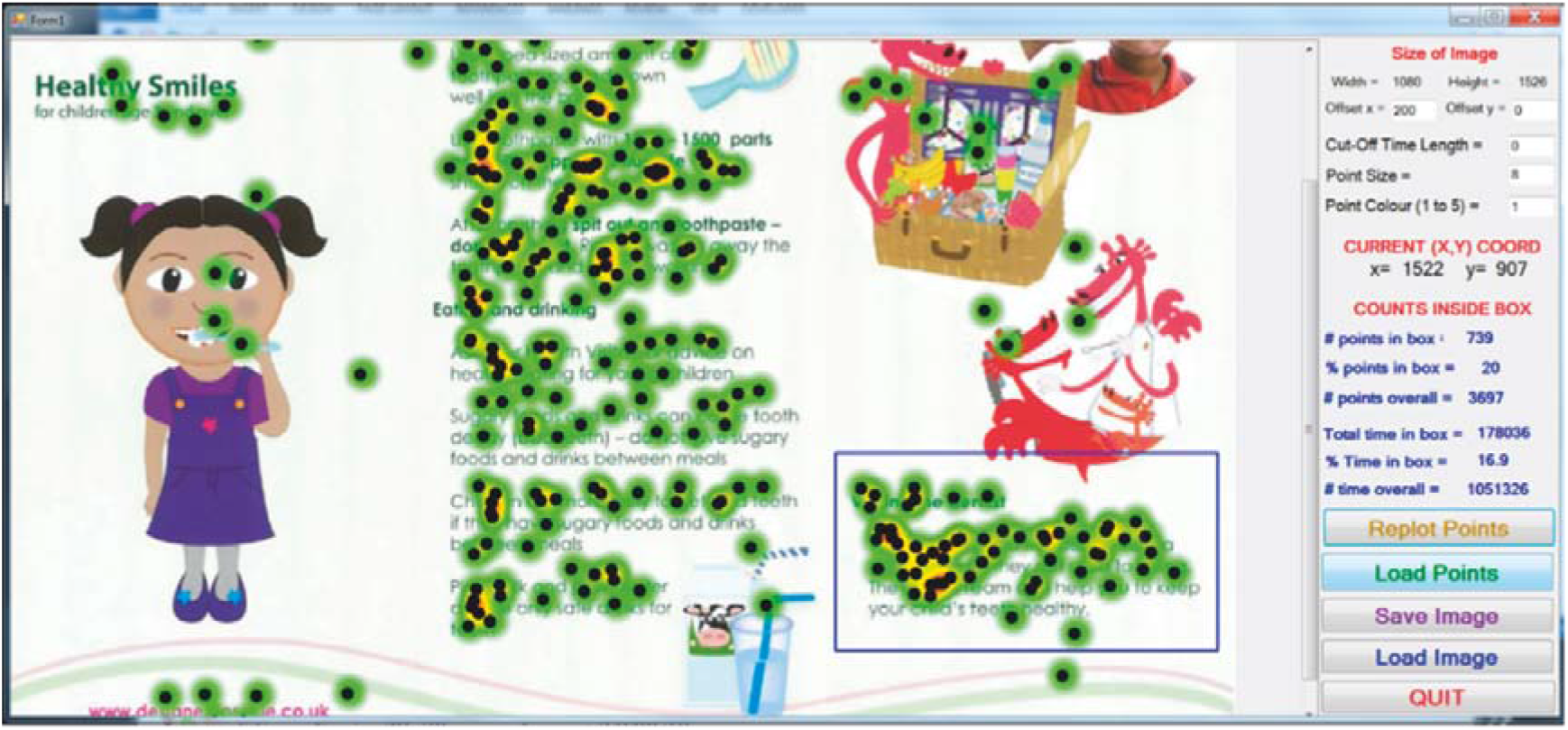
Graphical user interface created in Visual Basic to explore eye fixation data exported from Tobii Pro software as an EXCEL file (black circles / dots) and then superiomposed on a heat map produced by Tobii Pro directly.

#### Statistical Analysis

Survey results were transferred to Microsoft Excel and SPSS V27. Quantitative data was summarised using descriptive statistics for continuous variables (length of time spent) and ordinal data (overall rating of clarity of design). Frequency tables were used for the knowledge tests (yes/no). Statistical tests were carried out using SPSS V27 to investigate differences between the two PILs and potential confounding factors including sex, year of study, program of study, previous orthodontic treatment (orthodontic PIL only), parent or guardian (Designed to Smile PIL only), and whether the participant had seen the PIL previously. Generally, these tests were either non-parametric or chi-squared tests, as appropriate for the data type, and were carried out by a statistician. Statistical test type and associated results are quoted explicitly below. There was no missing data. The significance level for statistical tests was set at 0.05.

#### Thematic Analyses

Qualitative data from written free-text comments on the best design features of each PIL, suggested improvements to the PIL design, and any missing information from the PILs were analysed using word clouds and a simple thematic approach.

#### Data triangulation

Triangulation of survey data and eye-tracking data was carried out by “visual inspection” of heat maps and gaze plots. In this context, visual inspection means that we look at the heat maps and gaze plots and then try to see if there were any obvious differences in patterns between the highest and lowest scores by design and also between the highest and lowest quiz results for both PILs. We also wished to see if there were any obvious and large differences in the heat maps and gaze plots between the two PILs, e.g., relating to images and graphics, again through visual inspection.

## Results

### Participant Characteristics

53 participants were recruited in total (estimated 0.2% response rate based on a student cohort of 33,000 across the institution). Participant characteristics are shown in Table 1. All participants had good eyesight with or without corrective lenses.

**Table 1:** Participant characteristics. Percentages are formed with respect to a total sample size of 53 participants recruited to the study.

| **Programme of Learning** | **Law (or Law and Politics)** | **Mechanical Engineering** | **All Other Programmes ($n \leq 2$ in all cases)** |
| --- | --- | --- | --- |
| *n* (%) | 29 (54.7%) | 4 (7.6%) | 20 (37.7%) |
| **Biological Sex** | **Male** | **Female** |  |
| *n* (%) | 27 (50.9%) | 26 (49.1%) |  |
| **Orthodontic Treatment Previously?** | **No** | **Yes** |  |
| *n* (%) | 33 (62.3%) | 20 (37.7%). |  |
| **Are you a parent or guardian to any children?** | **No** | **Yes** |  |
| *n* (%) | 53 (100%) | 0 (0%) |  |
| **Have you seen this leaflet before? (Orthodontic Patient Information Leaflet)** | **No** | **Yes** |  |
| *n* (%) | 52 (98.1%) | 1 (1.9%) |  |
| **Have you seen this leaflet before? (Design to Smile Patient Information Leaflet)** | **No** | **Yes** |  |
| *n* (%) | 53 (100%) | 0 (0%) |  |

### Leaflet clarity

As shown in Table 2, 100% and 96.2% of participants responded that the PILs were either “clear” or “very clear” for the orthodontic PIL and the Designed to Smile PIL, respectively. However, 66% of participants (95% CI: 52.6% to 77.4%) responded that the Designed to Smile PIL was “very clear” compared to only 36.8% (95% CI: 24.3% to 49.3%) for the orthodontic PIL. The Designed to Smile PIL was therefore thought to be clearer than the orthodontic PIL (Wilcoxon signed-rank test: *P* = 0.006). Strong differences in ratings of clarity of design did not occur and all comparisons were not statistically significant (*P* > 0.05) with respect to course of study, year of study, sex, or previous orthodontic treatment. Only one participant out of fifty three had seen the orthodontic PIL before, none of the participants had seen the Designed to Smile PIL before, and none of the participants were parents or guardians to children. Statistical tests based on these factors were impossible therefore.

**Table 2:**
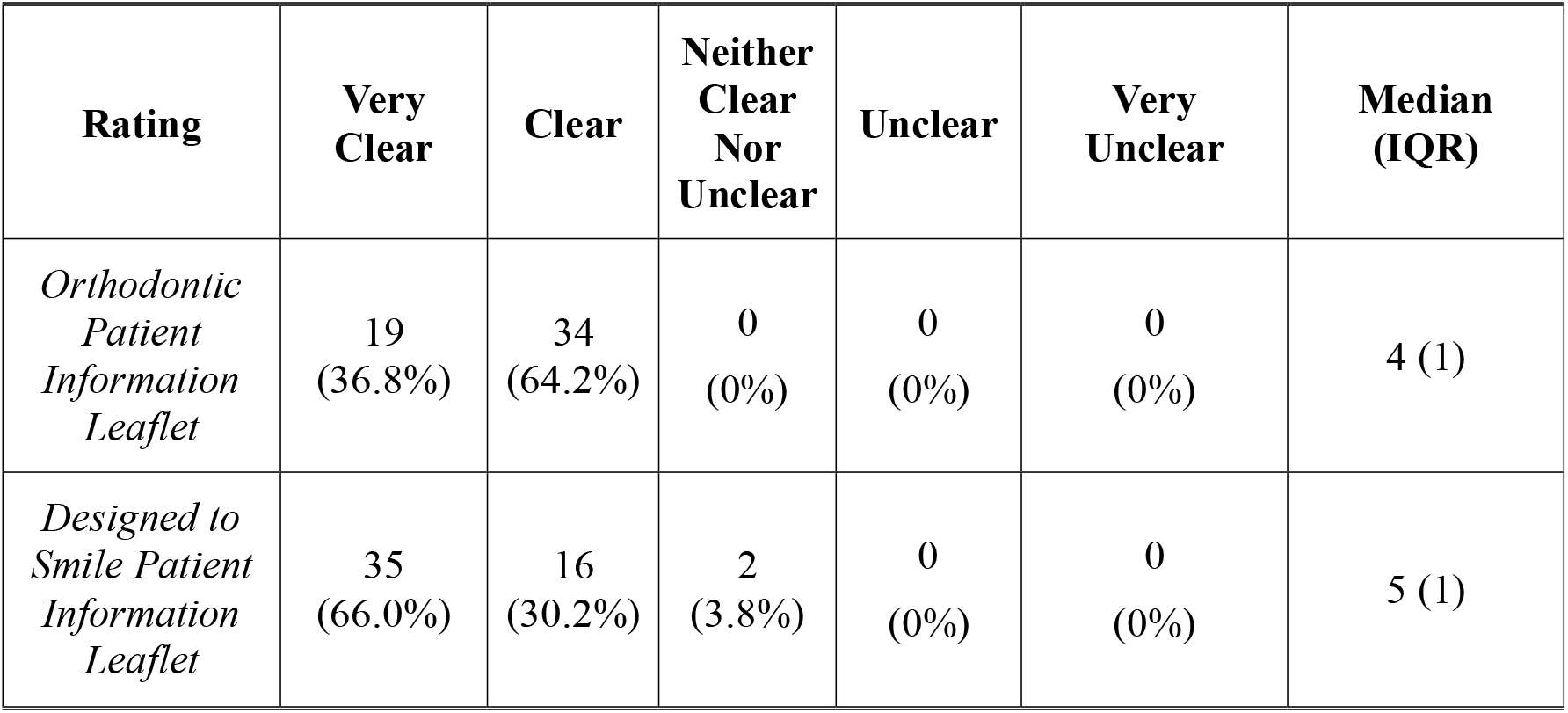
Number and percentage (in brackets) of participants responding to the question “Please rate how clear you thought the information leaflet was in the design survey”. Medians and inter-quartile ranges (IQR) of this ordinal variable were found by assigning a numerical value to the levels, i.e., 1 = very unclear, 2 = unclear, 3 = neither clear nor unclear, 4 = clear and 5 = very clear. The Designed to Smile PIL was found to be clearer than the BOS PIL (Wilcoxon signed-rank test: *P* = 0.006), shown by the fact that 66% of subjects responded that it was very clear compared to only 36.8% for the orthodontic leaflet.

| **Rating** | **Very Clear** | **Clear** | **Neither Clear Nor Unclear** | **Unclear** | **Very Unclear** | **Median (IQR)** |
| --- | --- | --- | --- | --- | --- | --- |
| *Orthodontic Patient Information Leaflet* | 19<br>(36.8%) | 34<br>(64.2%) | 0<br>(0%) | 0<br>(0%) | 0<br>(0%) | 4 (1) |
| *Designed to Smile Patient Information Leaflet* | 35<br>(66.0%) | 16<br>(30.2%) | 2<br>(3.8%) | 0<br>(0%) | 0<br>(0%) | 5 (1) |

### Opinions on the design of each leaflet

Across all free text comments, three main themes were found: design and readability, images and graphics, and content. Typical participant responses are provided in Table 3. Responses regarding the best features, areas for improvement, and any information participants felt were missing are discussed in more detail below.

**Table 3:** Some typical responses to the three text-based questions in the design survey, i.e., what participants thought were the best features, or suggestions as to how the leaflets could be improved, or what was missing, are quoted verbatim in this table. Participant identification numbers are shown also in the right-hand column, as well as which leaflet the comments refer to leaflet and in response which question in the survey. These comments are organised into global themes in the left-hand column, i.e.: Design & Readability, Images & Graphics, and Content.

| **Global Theme** | **Comment** | **Participant ID, Leaflet, & Question** |
| --- | --- | --- |
| Design & Readability | *It is laid out well and easy to follow, information is easy to understand* | ID = 3, Orthodontic, Best Features |
|  | *Simple and easy to the eye, not too much information leaving readers able to digest the information easily* | ID = 16, Designed To Smile, Best Features |
|  | *Important information are highlighted in bold making it clear to reader* | ID = 24, Designed To Smile, Best Features |
|  | *Information was well placed, there was an order in which understanding it was easy* | ID = 33, Designed To Smile, Best Features |
|  | *The points were split up into neat points. This allows us to pick up the info easily* | ID = 42, Designed To Smile, Best Features |
|  | *There are too many words. I appreciate the detail but it was overloaded with too much information on just two pages* | ID = 29, Orthodontic, Suggested Improvements |
| Images & Graphics | *It included illustrations and pictures which helped with my understanding, it was very detailed and easy to understand* | ID = 29, Orthodontic, Best Features |
|  | *Images would help kids attention and easy to understand* | ID = 24, Designed To Smile, Best Features |
|  | *As someone who appreciate creativity, leaflet should be improved in terms of designing. More infographics and well designed* | ID = 20, Orthodontic, Suggested Improvements |
|  | *Maybe more pictures, I am a very visual person and I find easier to understand information if it is presented in a visual form, too much text will bore me* | ID = 31, Orthodontic, Suggested Improvements |
|  | *By including more photo and diagrams rather than more words* | ID = 40, Orthodontic, Suggested Improvements |
| Content | *Some review from people would be great as an insight to show the importance of healthy teeth* | ID = 11, Orthodontic, What was missing |
|  | *Public feedback or review from patients-maybe before and after of using braces* | ID = 35, Orthodontic,<br>What was missing |

### Best features of each leaflet

Comments with regards to the “best features” of the two PILs are summarised as word clouds in Figure 4. Participants found both PILs easy to understand, as shown by specific comments in Table 3 under the global theme of “design and readability”. Similarly, the use of graphics or images was strongly appreciated by participants and often enhanced understanding, especially for the more complex orthodontic PIL, as shown by specific comments in Table 3 under the global theme of “images and graphics”.

**Figure 4:**
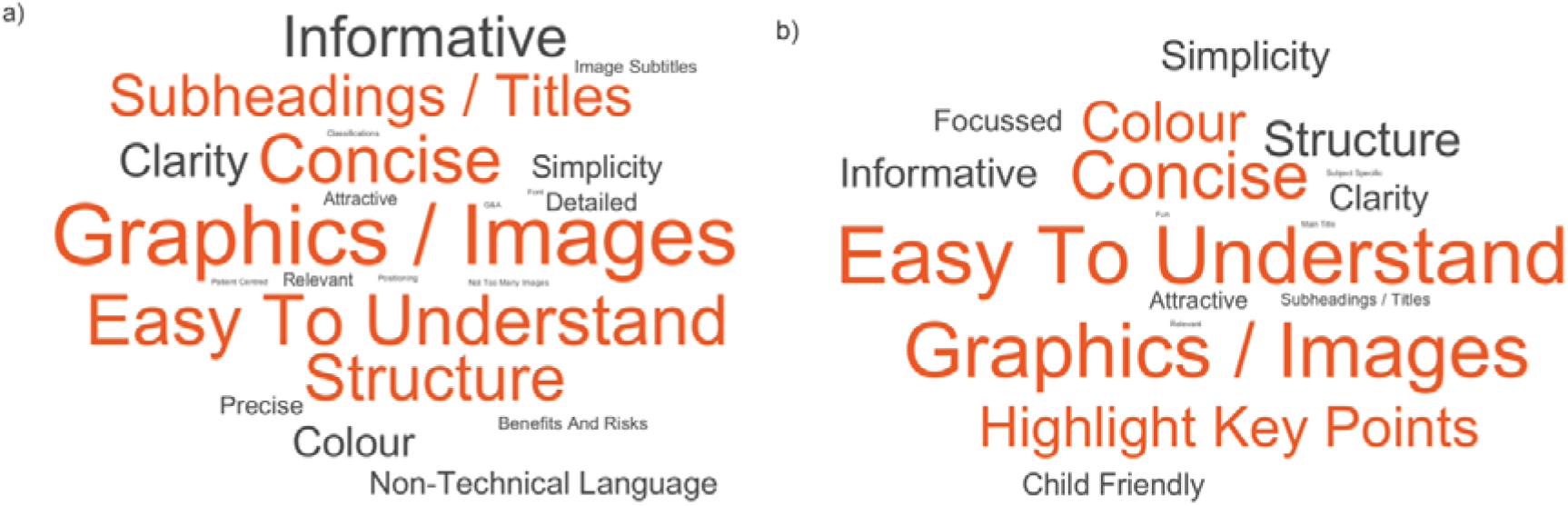
Word clouds for qualitative themes found for the two leaflets on the “Best Features”: (a) British Orthodontic Society Patient Information Leaflet; (b) Designed to Smile Participant Information Leaflet. The font size of the words in the figures indicates the number of comments belonging to each of these themes.

The child-friendliness of the Designed to Smile PIL was also noted by some participants, which is logical as Designed to Smile is aimed at improving oral health in primary school children. Participants appreciated efforts to make the PILs concise, clear, and simple, e.g., non-technical wording, for the orthodontic PIL where possible; this is shown clearly in the themes shown in Figure 4. Anything that helped the readers navigate the PILs or to recognise the importance of the text was also appreciated, including the use of subheadings or titles for specific sections or if key points were highlighted in some manner (e.g., use of bold font or colour). As shown by comments in Table 3, the readability of both PILs was enhanced by use of a logical structure, including figures or images.

### Suggestions for improvement in leaflet design

The free text comments relating to suggestions for improvements are summarised as word clouds in Figure 5. Notably, 20 out of 53 participants (37.7%) either did not respond or said “none” or “no” in text-based comments for the Designed to Smile PIL compared to 7 (13.2%) participants for the orthodontic PIL (Binomial / Sign test: *P* = 0.002). This further supports the result that the Designed to Smile PIL was more highly rated for clarity of design than the orthodontic PIL.

**Figure 5:**
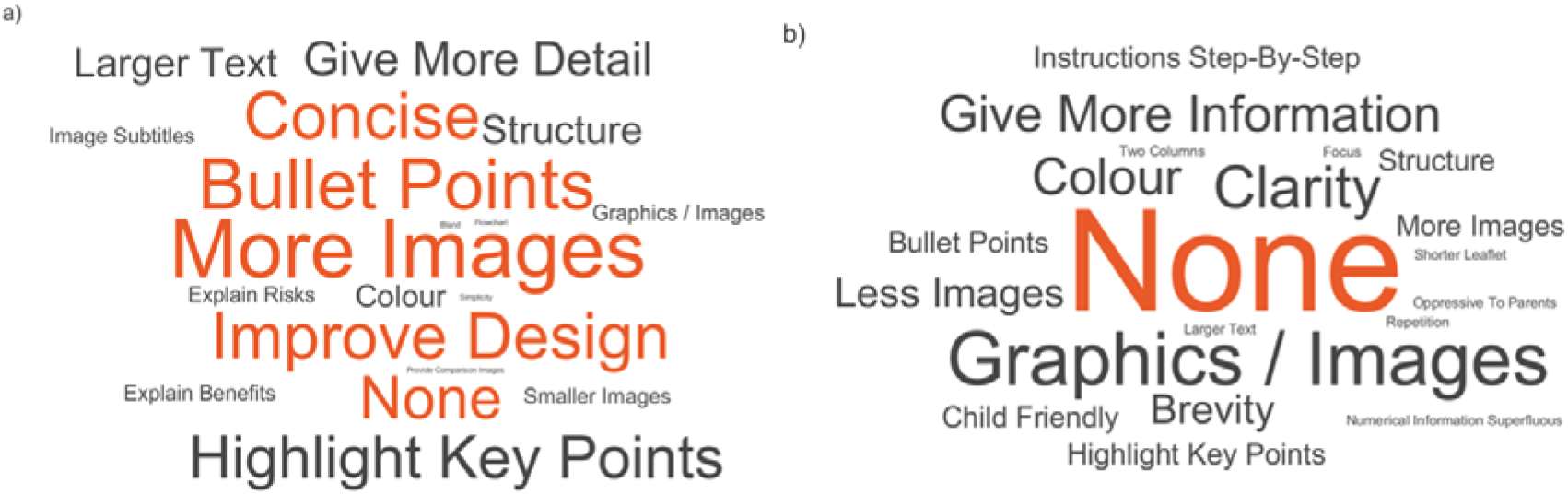
Word clouds for qualitative themes found for the two leaflets for the topic “Suggestions for Improvements”: (a) British Orthodontic Society Patient Information Leaflet; (b) Designed to Smile Participant Information Leaflet. The font size of the words in the figures indicates the number of comments belonging to each of these themes.

Comments for the orthodontic PIL tended to reflect the more difficult content, problems with structure and presentation, and the need to convey a larger amount of information as blocks of text. The use of images rather than text was mentioned by several participants, as shown by specific comments in Table 3 under the global theme of “images and graphics”. The comments in Table 3 also suggest that overall design of the orthodontic PIL was an issue for participants with a desire for the PIL to be more concise. The use of bullet points or highlighting key points to improve readability was suggested. Overall, there were more comments suggesting improvements for the orthodontic PIL than for the Designed to Smile PIL.

### Missing information

Themes were also identified from free-text comments for the subject of “missing information,” where comments are summarised as word clouds in Figure 6. 43 out of 53 participants (81.1%) either did not respond or said “none” or “no” for the Designed to Smile PIL when asked if there was any missing information, whereas 35 participants (66.0%) provided suggestions for the orthodontic PIL (Binomial / Sign test: *P* = 0.039). Missing information was specific to the topics in each PIL. Common suggestions related to the wish for more details or specific information, particularly relating to risks of orthodontic treatment, but also the potential benefits and duration of treatment. For the Designed to Smile PIL, improvements related to aspects of maintaining good oral health, e.g., duration of tooth-brushing or regularity of visiting the dentist. Interestingly, two participants asked for a description of the patient experience to be added to the orthodontic PIL, as shown by two comments in Table 3 under the global theme of “content”.

**Figure 6:**
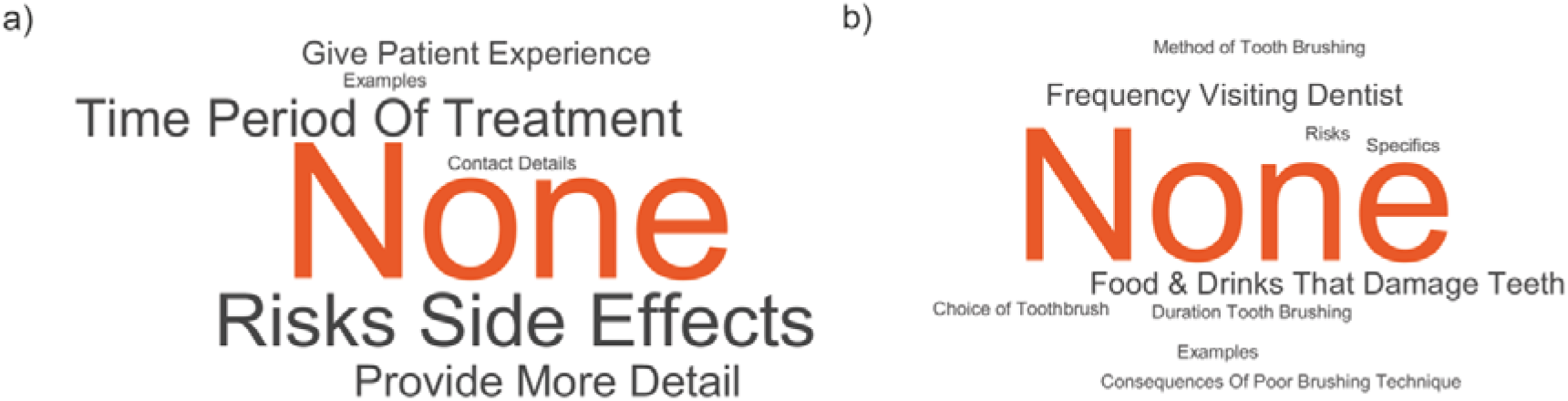
Word clouds for qualitative themes found for the two leaflets for the topic “Missing Information”: (a) British Orthodontic Society Patient Information Leaflet; (b) Designed to Smile Participant Information Leaflet. The font size of the words in the figures indicates the number of comments belonging to each of these themes.

### Results of Eye Tracking

Results for heat maps eye gaze patterns are shown in Figures 7 and 8 for both PILs. Results are shown for those participants who obtained the maximum and minimum quiz scores and similarly also ratings of clarity of design, i.e., extremes of both measures to see if there are any differences in these patterns. Results form gaze plots indicated that participants followed the order of the text, i.e., broadly top left to bottom right, although they did also look at the images and some participants reread some sections of the text, which altered this flow also.

**Figure 7:**
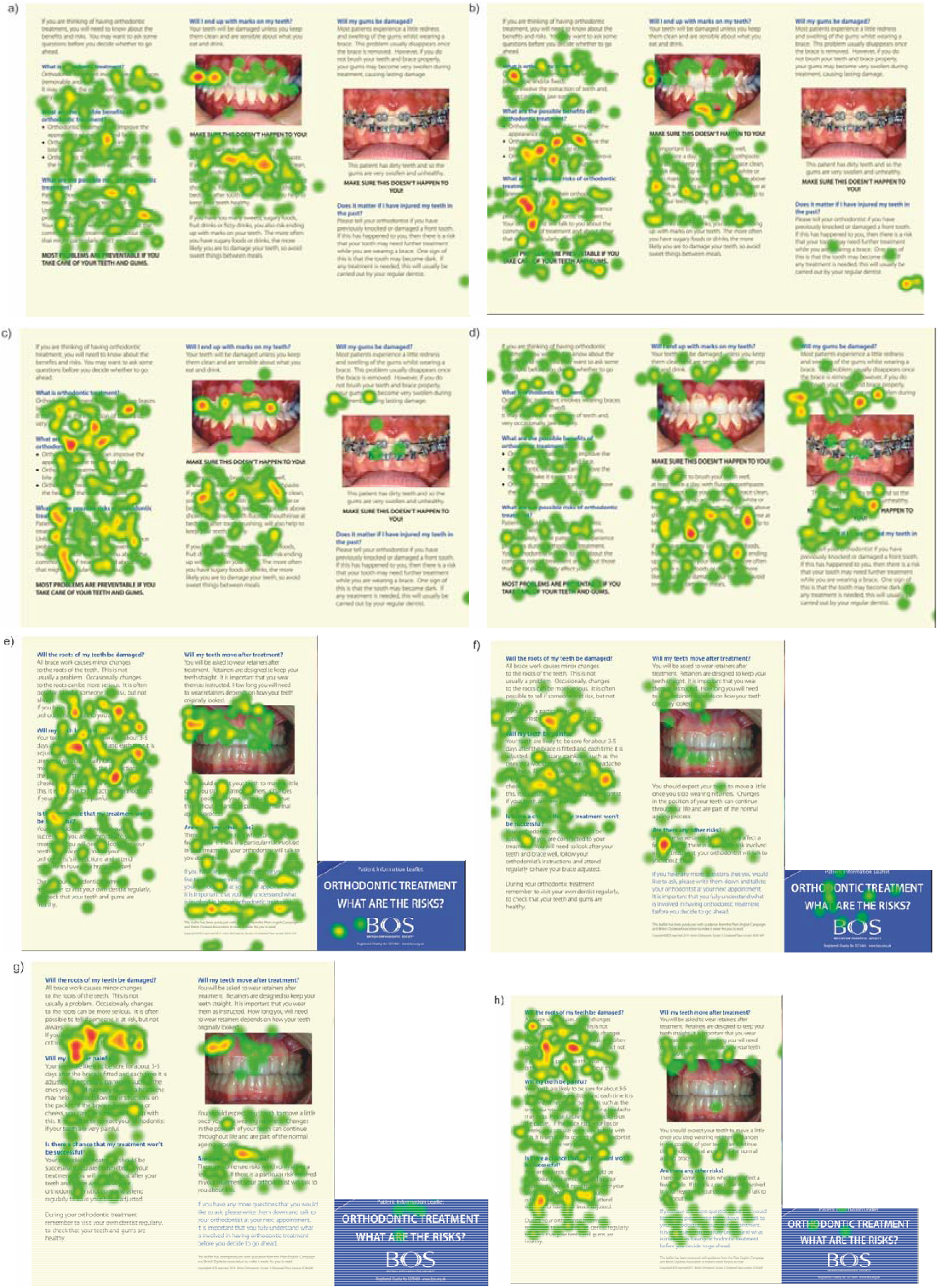
Heat map of eye gaze patterns from eye tracking for page 1 of the orthodontic leaflet. Green then yellow and then red colours indicate increasing amounts of time spent looking at specific features in the image. (page 1 = image a and page 2 = image d) Participant ID = 19 (quiz score = 35%; rating of design = very clear; this Participant thought the leaflet could have been improved by inclusion of more images in free-text comments); (page 1 = image b and page 2 = image e) Participant ID = 20 (quiz score = 35%; rating of design = neither clear not unclear; this Participant stated that images were a best feature in free-text comments); (page 1 = image c and page 2 = image f) Participant ID = 28 (quiz score = 60%; rating of design = clear; this Participant thought there were not too many images int the leaflet, but that they could be smaller in size in free-text comments); (page 1 = image d and page 2 = image g) Participant ID = 29 (quiz score = 60%; rating of design = very clear; this Participant thought that “illustrations and pictures which helped with my understanding” in free-text comments).

**Figure 8:**
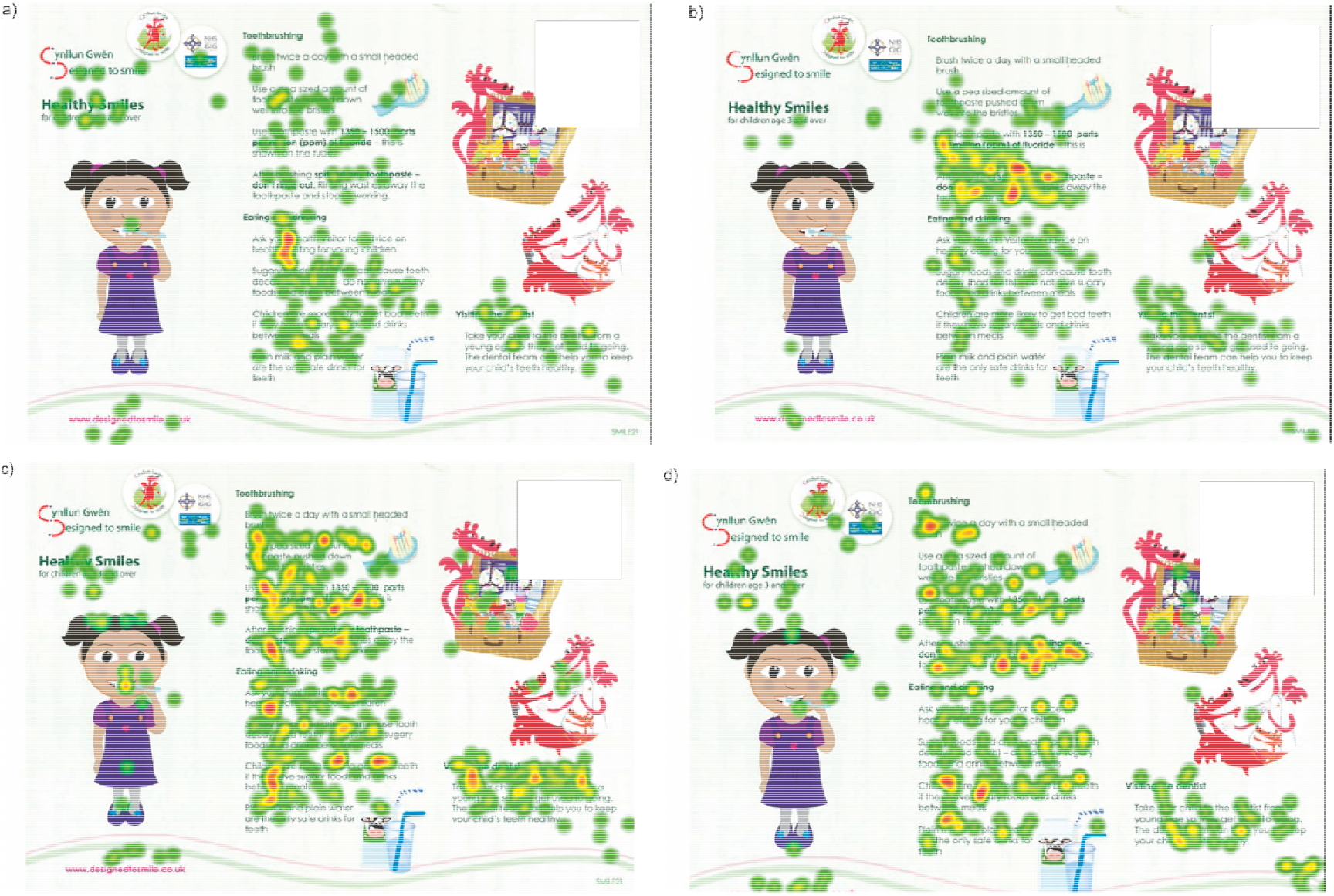
Heat map of eye gaze patterns from eye tracking for the Designed to Smile information leaflet. Green then yellow and then red colours indicate increasing amounts of time spent looking at specific features in the image. (a) Participant ID = 7 (quiz score = 60%; ratings of clarity of design = very clear; this participant made no comments about images and thought that the leaflet could not be improved in free-text comments); (b) Participant ID = 19 (quiz score = 60%; ratings of clarity of design = very clear; this participant thought the leaflet could have been improved by inclusion of clear infographics in free-text comments); (c) Participant ID = 29 (quiz score = 100%; ratings of clarity of design = very clear; this participant made no comments about images and thought that the leaflet was easy to understand in free-text comments); (d) Participant ID = 50 (quiz score = 100%; ratings of clarity of design = clear; this participant made also no comments about images and thought that the leaflet was easy to understand in free-text comments).

Results from the eye-tracking were also analysed quantitatively by using the graphical user interface shown in Figure 3 to measure how much time each participant spent looking at specific areas of each PIL. Ratings of clarity of design of the PILs and quiz results were not strongly correlated (*P* > 0.05 for all statistical tests) with overall fixation times or specific fixation times looking specific elements of the PILs measured by the quiz or even if they looked specifically at these elements at all or not. By looking at Figures 7 and 8, we see that there are no obvious or large differences in the heat maps between the highest and lowest scores by design and between the highest and lowest quiz results.

Eye tracking also indicated that participants spent a mean average of 24.6% (SD = 17.7%; median = 22.2%) of their time on average looking at graphics or images for the orthodontic PIL, compared to a mean of 9.6% (SD = 8.5%; median=8.2%) for the Designed to Smile PIL, i.e., a factor of almost two-and-a-half times longer for the orthodontic PIL compared to the Designed to Smile PIL (Wilcoxon signed-rank test: *P* < 0.001). This can also be seen by visual inspection of Figures 7 and 8, where there is much more “red colour” in the heat maps superimposed on the images for the orthodontic PIL compared to the heat maps superimposed on images and graphics in the Designed to Smile PIL.

### Results for Quizzes Measuring Knowledge Retention

The mean percentage of correct answers across all items in the orthodontic PIL was 48.9% (SD = 9.0). The mean percentage of questions that were answered correctly for the Designed to Smile PIL was 80.0% (SD = 11.3%), which was significantly higher than for the orthodontic PIL (Wilcoxon signed-rank test: *P* < 0.001). Percentages of correct answers and ratings of clarity of design for PILs 1 and 2 were not strongly and generally not significantly different by course of study and by sex (Mann-Whitney test: *P* > 0.05). Previous orthodontic treatment did seem to lead to improved chances of responding correctly slightly in the orthodontic PIL, although results were often not significant (Fisher’s exact test: *P* > 0.05) and so this effect could have happened by chance.

## Discussion

### Interpretation of Main Findings

An important result found here was that 100% and 96.2% of participants responded that the PILs were either “clear” or “very clear” for the orthodontic PIL and the Designed to Smile PIL, respectively. Therefore, both PILs were acceptable to participants. Indeed, many of the aspects of good PIL design seen in the literature were echoed strongly in free-text comments for both PILs, including use of images, text font and colour, simplicity of language, highlighting key points, and brevity(Kitching, 1990; Dowse *et al*., 2011; Adepu *et al*., 2012; Brotherstone *et al*., 2006; Jansen *et al*., 2008; Karreman *et al*., 2007; Pires *et al*., 2015; Pothier *et al*., 2008). However, some participants noted that the design of the orthodontic PIL could be improved in free-text comments, e.g., by the use of highlighting key points, breaking up large blocks of text, or possibly even more use of informative images or graphics. An interesting comment by two participants was that a description of the patient experience for the orthodontic PIL would be beneficial.

Knowledge after reading the PILs was significantly (*P* < 0.001) higher for the Designed to Smile PIL (80% correct) compared to the orthodontic PIL (48.9% correct). Differences in knowledge after reading the PILs may be explained by the design of the PIL but may also be influenced by other factors. These factors include: the more complex orthodontic information compared to the information in the Designed to Smile PIL; the time participants were given to read the PILs (75 seconds per page); and that information in the Designed to Smile PIL is more likely to be common knowledge to the general public. These factors go some way to explaining why knowledge after reading the PILs was significantly higher for the Designed to Smile PIL compared to the orthodontic PIL and a pre-test knowledge test would have allowed this to be assessed. However, the results confirm that generally, participants have gaps in their knowledge of orthodontic risks after reading the PIL for 75 seconds per page. This suggests improvements could be made to the PIL content to improve knowledge transfer, which is supported by the free-text comments provided.

Furthermore, eye tracking indicated that no strong differences in knowledge after reading the PIL were observed as a function of time looking the specific elements of knowledge in the PILs being tested. Participants who looked at the relevant information in the orthodontic PIL did not retain any more knowledge than those participants who did not, i.e., key pieces of orthodontic information need to come through much more clearly, which is an important finding of this research.

Suggestive results for the use of images in the PILs was also found via eye tracking and text-based comments. This is also perhaps where eye tracking showed the most promise as it provided hard evidence relating to how much time was spent by participants on the images for both PILs. Very strong differences were seen in terms of length of time spent looking at images, which was much larger for the orthodontic PIL than for the Designed to Smile PIL. Indeed, a key theme noted by participants in free-text comments was that images were really useful in the orthodontic PIL to convey factual information. By contrast, they were generally thought less useful from free-text comments about the Designed to Smile PIL, where they were used more as graphics to attract the eye and / or to make the PIL more child friendly. Please note that text-based comments from participants did not indicate that this was because there was more content to cover for the orthodontic PIL, although it was clearly the more technically demanding of the two PILs and so images were found to be useful to illustrate these points.

The absolute time looking at specific elements of the PILs was measured here via eye tracking and the graphical user interface. This analysis gave an almost identical pattern of results to those results for the percentage time.

### Implications for Clinical Practice

We recommend that the insights presented above be used to improve the design of the orthodontic PIL to improve knowledge transfer (Patel *et al*., 2008; Thickett *et al*., 2006, Brotherstone *et al*., 2006; Meppelink *et al*., 2015), although we remark again that both PILs were generally well-received in terms of clarity of design.

### Limitations

A problem of eye-tracking is that it can be hard to synthesise and understand the detailed time-series data that it provides. Eye-tracking also creates massive amounts of information, which can therefore be hard to analyse. Here we used a graphical user interface that was very useful in providing “hard” quantitative evidence for the eye-tracking data, which will be used in future studies as it is simple and straightforward to use. However, this approach is also inefficient and is somewhat crude as it requires a lot of time-consuming manual intervention to extract numerical evidence for specific regions of interest only that can be selected by the user. Undoubtedly, specialised mathematical and / or AI tools are necessary to extract the most from eye-tracking data in future.

The use of two PILs by the same set of participants gave a deeper insight into design as it was possible to compare results readily; this was another strong advantage of the research presented here, i.e., we could work out what worked well and what did not, e.g., the relative simplicity of design for the Designed to Smile PIL compared to the orthodontic PIL.

Finally, each participant was asked to look at each image for 75 seconds without any further instructions, sample sizes were slightly low, i.e., 53 participants, and no base line knowledge was recorded prior to viewing the PILs. 75 seconds per page might not have been long enough for adequate retention of this information. These are further limitations of the research design in this initial exploration of orthodontic PIL design.

### Generalisability

An undergraduate student population was recruited, excluding those studying a healthcare-related subject, as a proxy for a patient population, albeit drawn from a population of students at Cardiff University. However, there are issues of external validity as a university population is likely to be more highly educated than the general UK population on average and the sample recruited does not reflect the large age range of potential readers of either PIL. Moreover, the method of recruitment does not account for non-response bias, with the participants actively consenting to participate in the study. Further work should explore a patient cohort for the orthodontic PIL and an appropriate cohort of parents / children for the Designed to Smile PIL.

### Implications for Further Research

Future studies will also use verbal cues to improve the sensitivity and specificity of the eye-tracking data collected, e.g., “please look at the text / images in this image” or “please look at the things in this image you feel are good / bad”. Varying the time spent viewing images is another obvious refinement in future and larger sample sizes will be used. Further studies could also focus on alternative media, including interactive digital information PILs. Future research might also address improvements in knowledge by using and pre-to post-design and how changes in design affect results.

## Conclusions

The purpose of this study was to evaluate an orthodontic information PIL that is commonly used in the UK and to compare it to a simpler PIL that encourages children to adopt good oral health habits as part of the Designed to Smile campaign in Wales. An aim of the study was to carry out quantitative ratings of the clarity of the design, which indicated that participants thought that both PILs were either clear or very clear. Another aim was to gather qualitative evidence relating to design and content via free-text comments. These comments indicated that improvements in design for the orthodontic PIL in particular could be made by highlighting key points, breaking up large blocks of text, increased use of informative images, and including descriptions of the patient experience. The literature in the field of patient information improving design and readability should improve knowledge retention. A final primary aim was to measure knowledge retention via quizzes, which showed that this was much lower for the orthodontic PIL compared to the simpler the Designed to Smile PIL measured via a quiz after reading each PIL. A secondary aim was to carry out eye tracking as participants read the PILs. Eye tracking provided a wealth of detailed information that proven challenging to analyse, although it supported comments of participants that images were very useful in explaining technical details to readers for the orthodontic PIL in particular.

## Data Availability

Data for this study available upon reasonable request to the authors.

## Appendix A

The Tobii Pro Nano shown in Fig. 1 is a screen-based eye tracker which captures eye-tracking data at 60Hz and is designed for fixation-based studies. The eye tracker was placed below the screen of a Dell Latitude 5490 laptop. Note that Tobii Pro Nano comes with Tobii Pro Lab software that allows image capture and data analysis, including gaze plots and heat maps. This software allows also one to export the eye-tracking data to an MS EXCEL file as Cartesian “gaze point” data. Data collection was carried out an orthodontic information leaflet (see Fig. 2a,b), as well as a simpler “Designed To Smile” information leaflet (see Fig. 2c) in order to provide a useful comparison. Eye-tracking involves the following elements. Firstly, the participant was seated comfortably in front of the laptop and the correct eye position was established. A standard calibration step was carried out each time a participant used the eye-tracker for recording. During calibration, the participant was asked to look at specific calibration dots that appear at multiple locations on the plane where the stimulus is located. The Tobii Pro software then corrected for any potential drift after calibration. This vital step removed biases / measurement error from the eye-tracking data.

## Appendix B

Black dots in Fig. 3 indicate fixation points from a single user for data extracted from MS EXCEL file for the Designed to Smile leaflet. These black dots are superimposed on a heat map of fixation gaze points obtained directly from the Tobii Pro Lab sofware and one sees that the dots match perfectly with the areas of high fixation from the Tobii Pro Lab software, shown in yellow and green. This is an excellent quantitative test of the graphical user interface. Specific features are selected by the user by using the blue box, which can be set using the mouse and “left-clicking” appropriately. The number (and percentage) of fixation points and total length number (and percentage) of time within the box can found, and results are given by the text on the right-hand side of the figure. In this manner, hard numerical evidence could be extracted from the eye-tracking data relating to specific features in the image.

## Appendix C

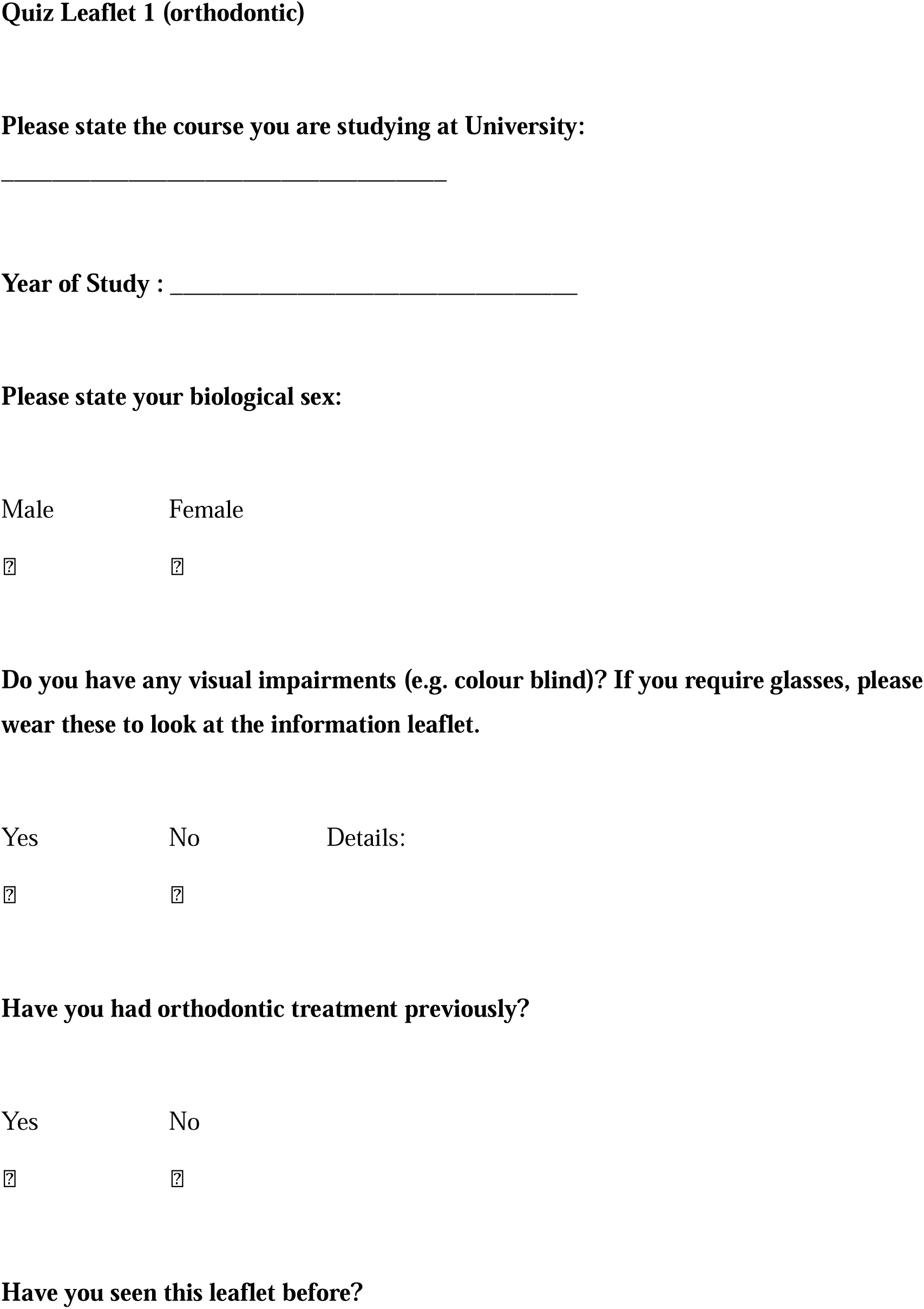

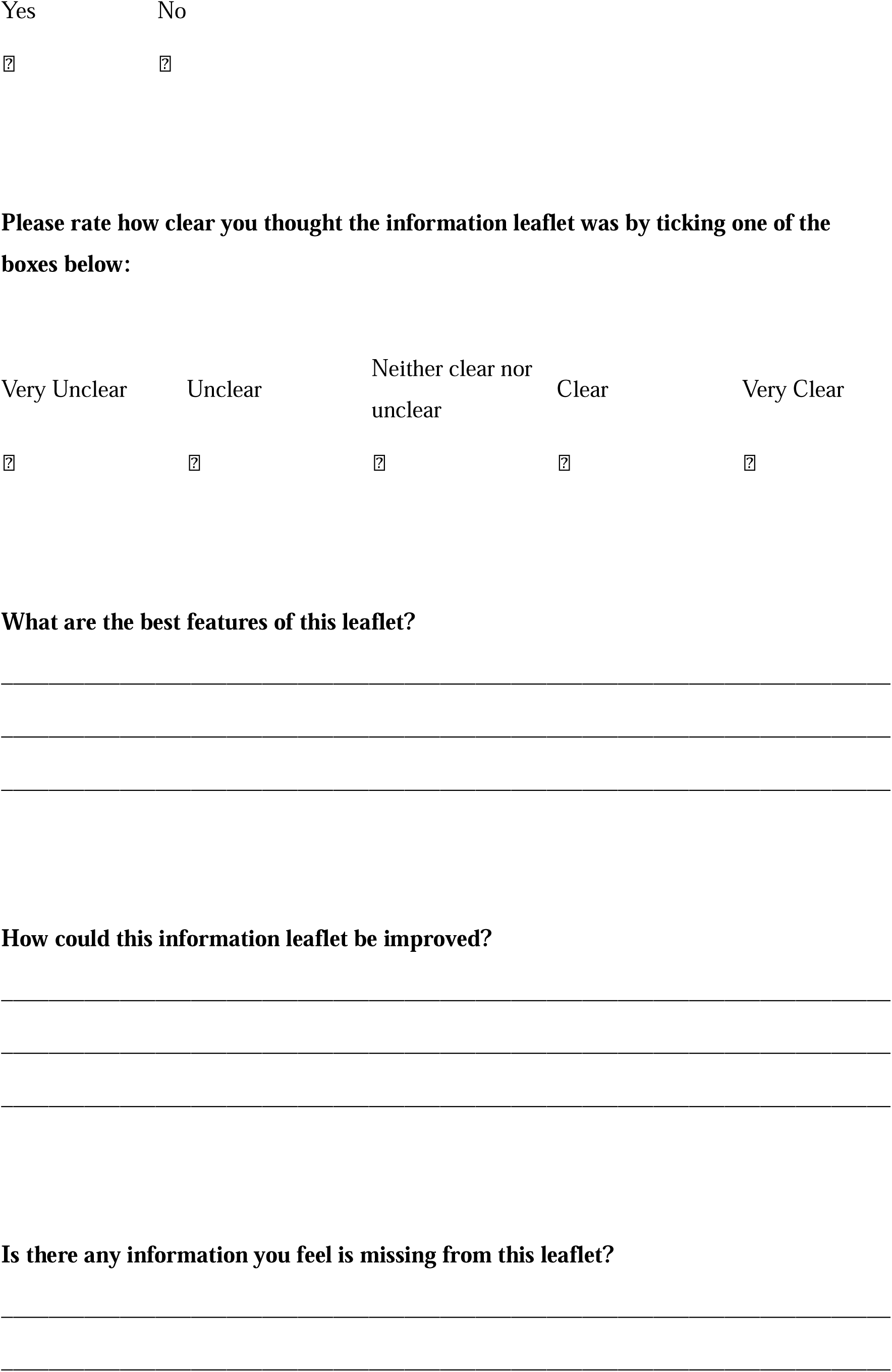

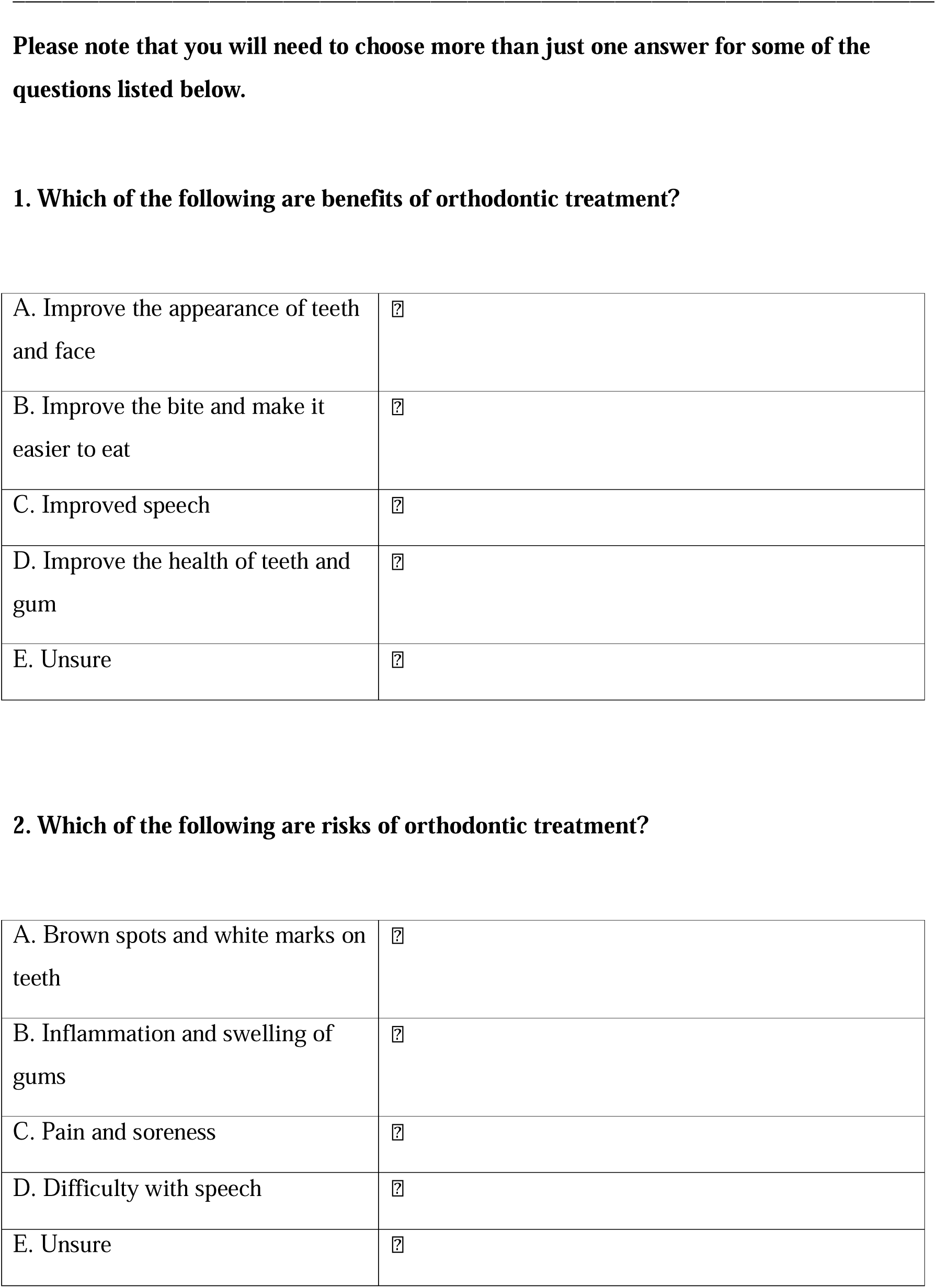

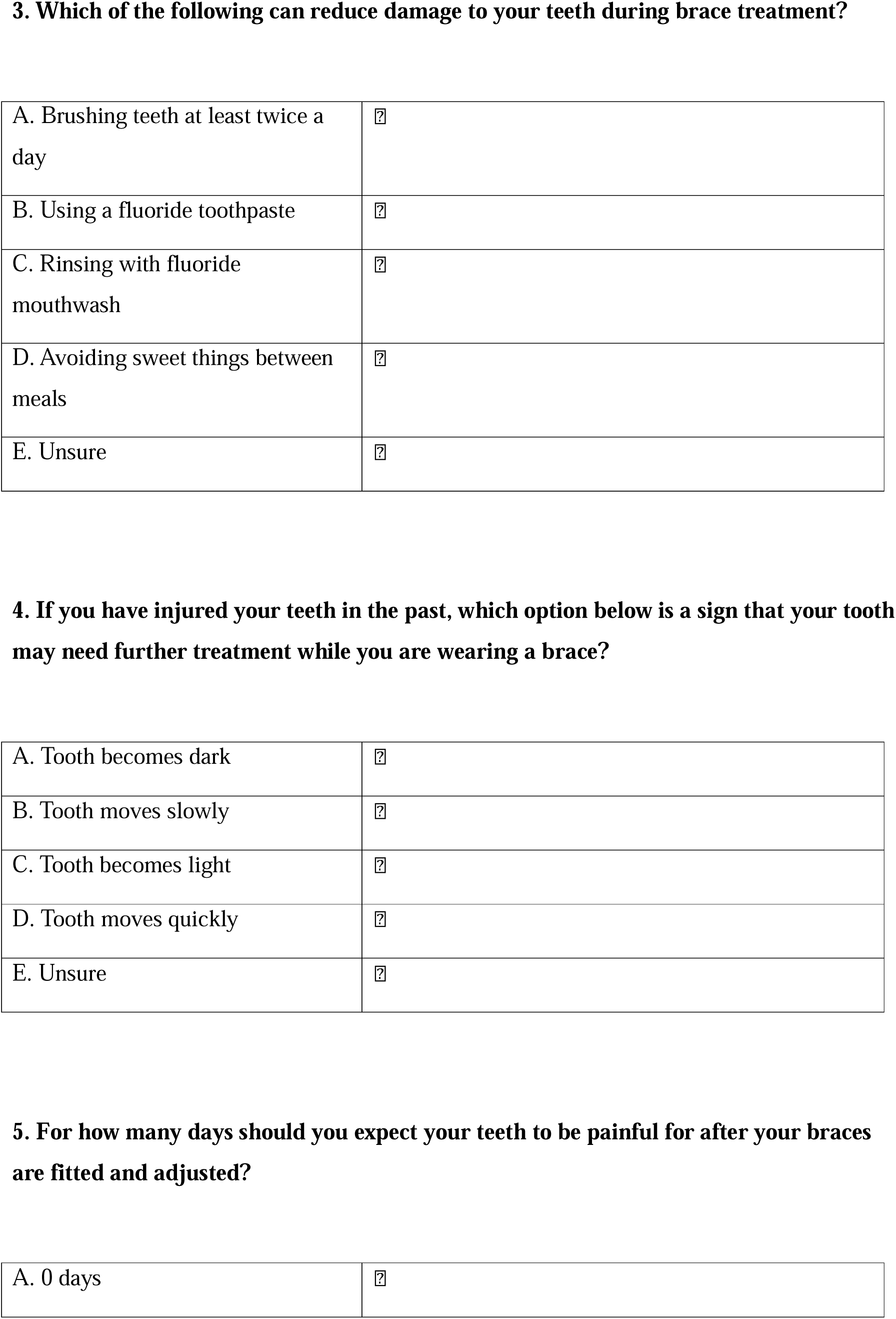

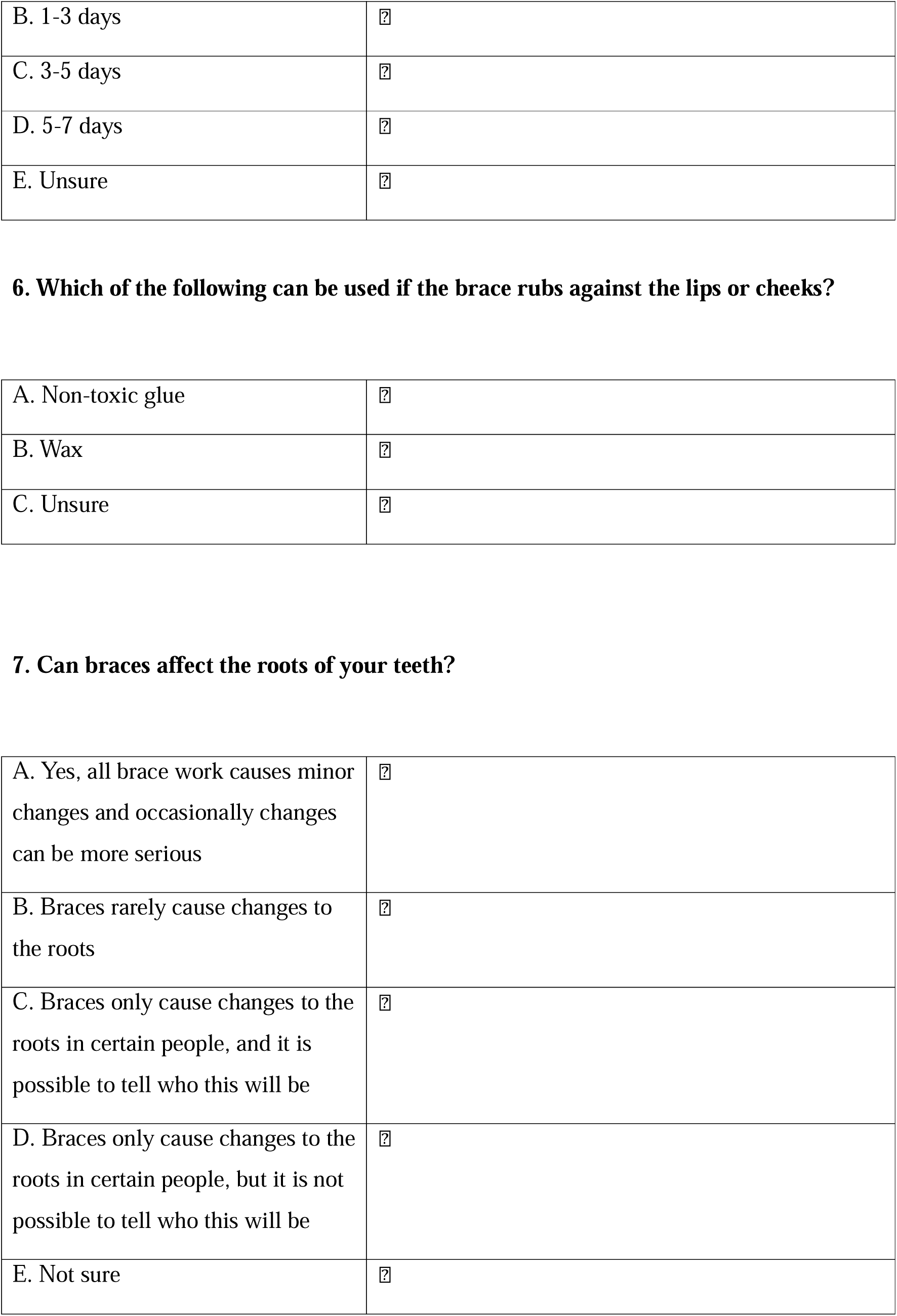

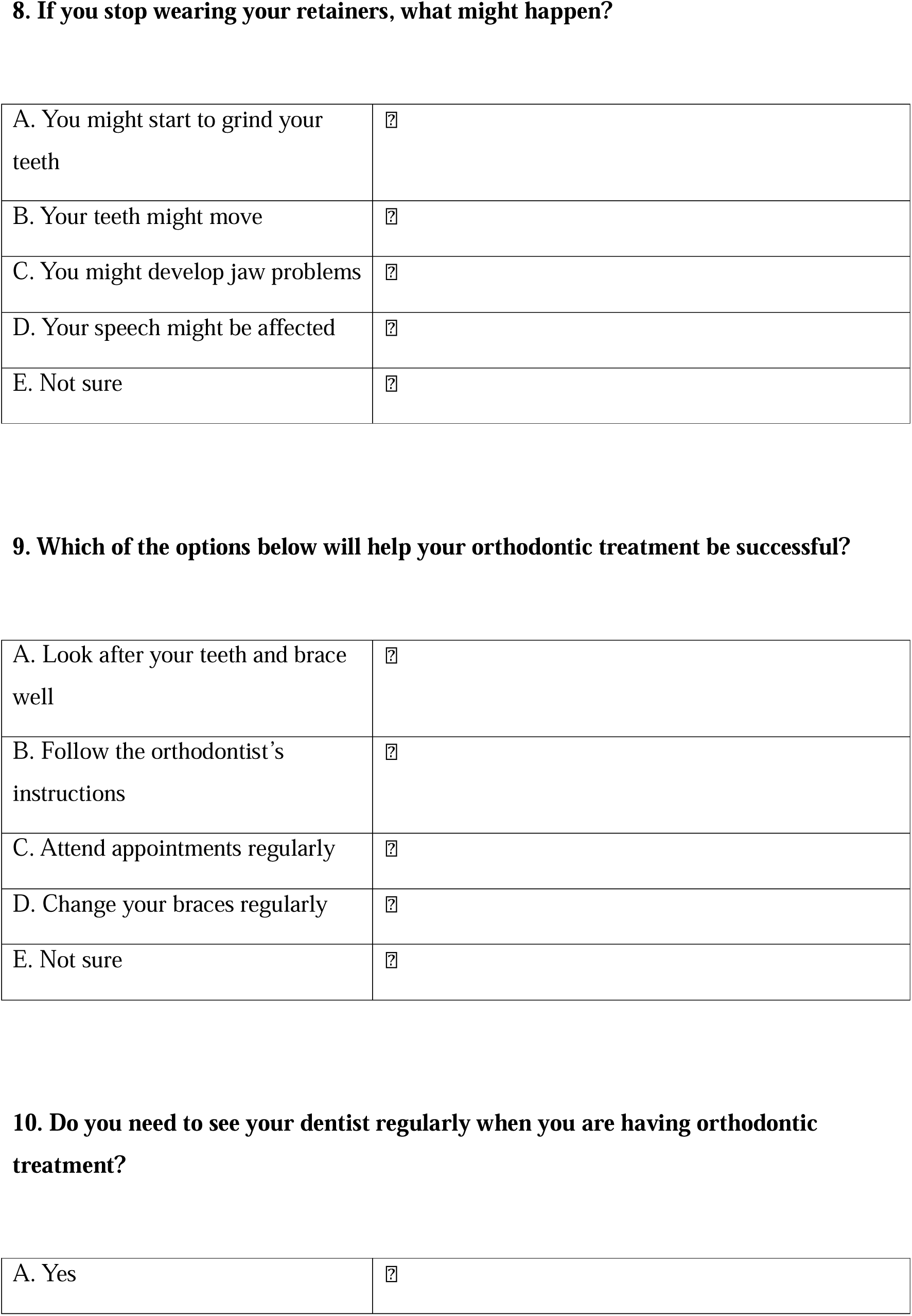

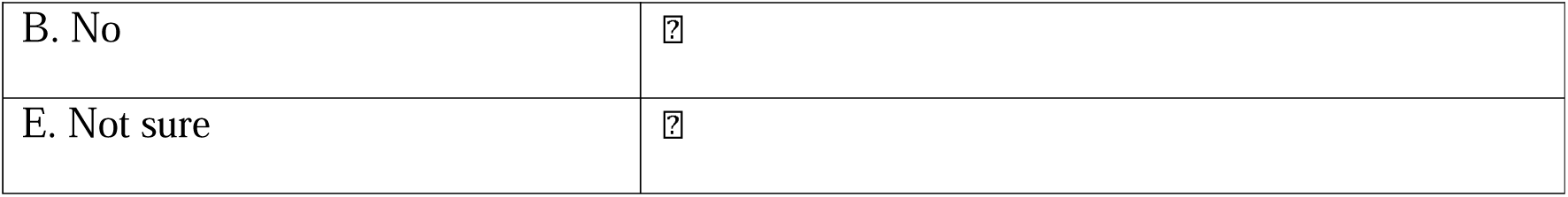

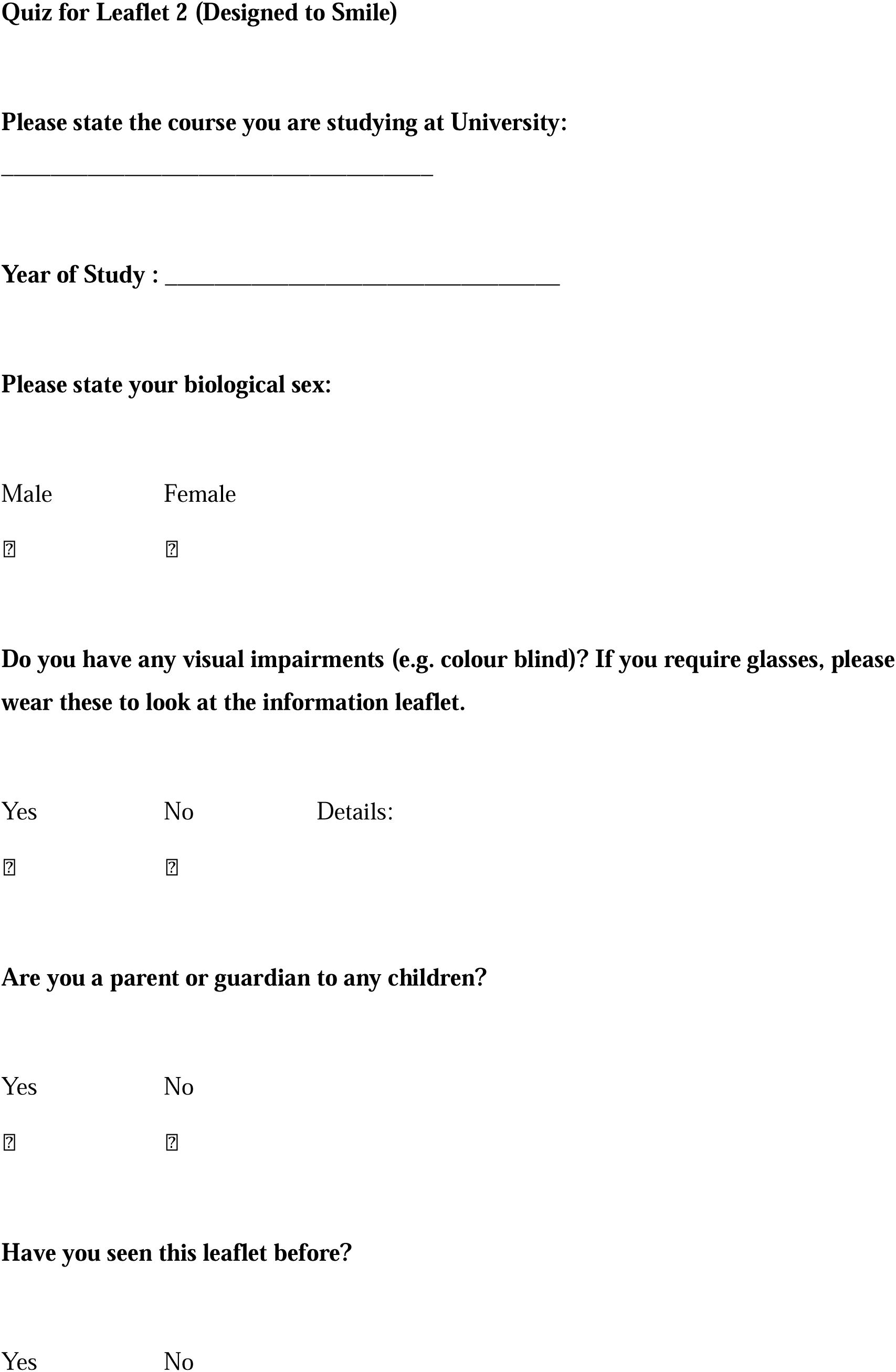

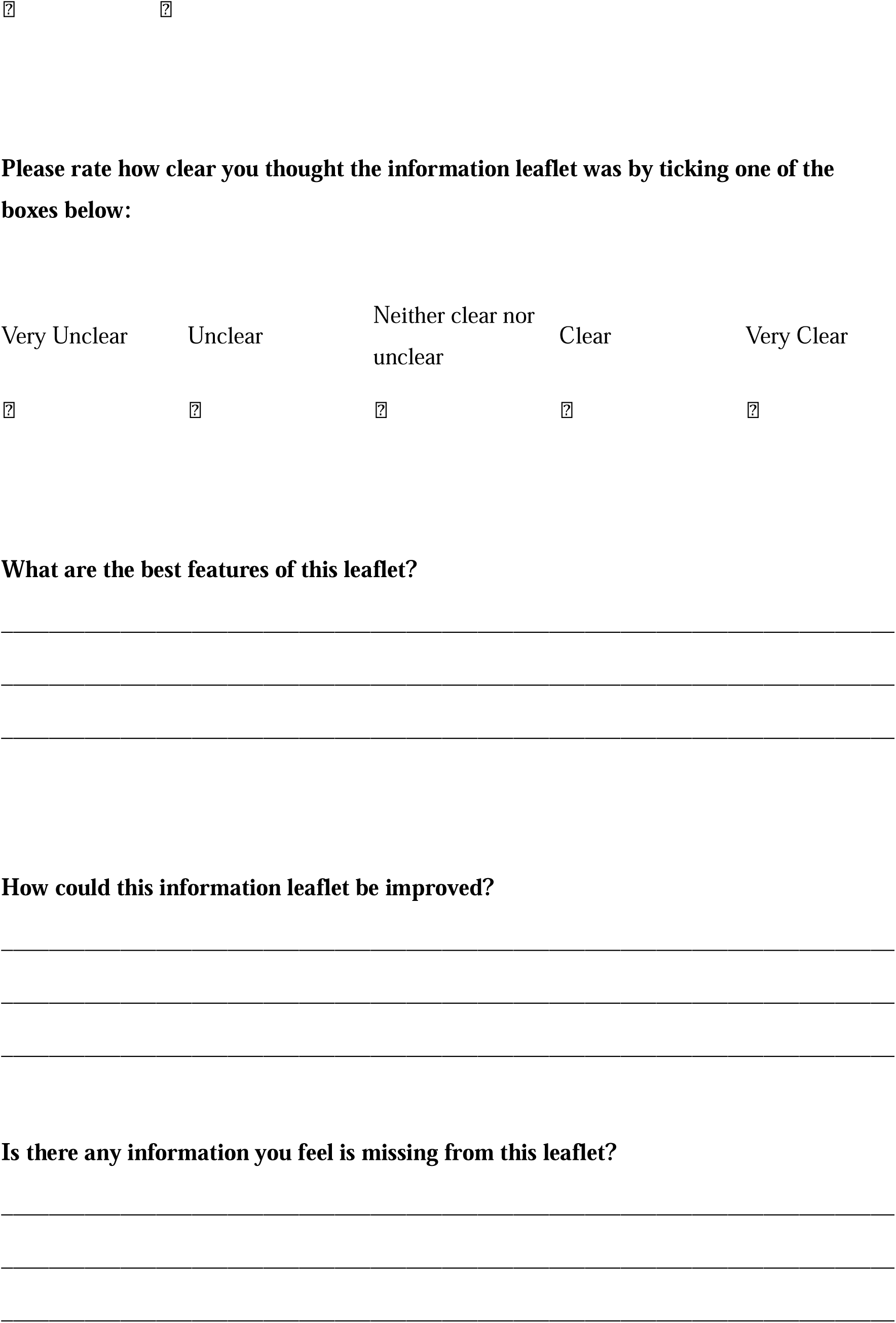

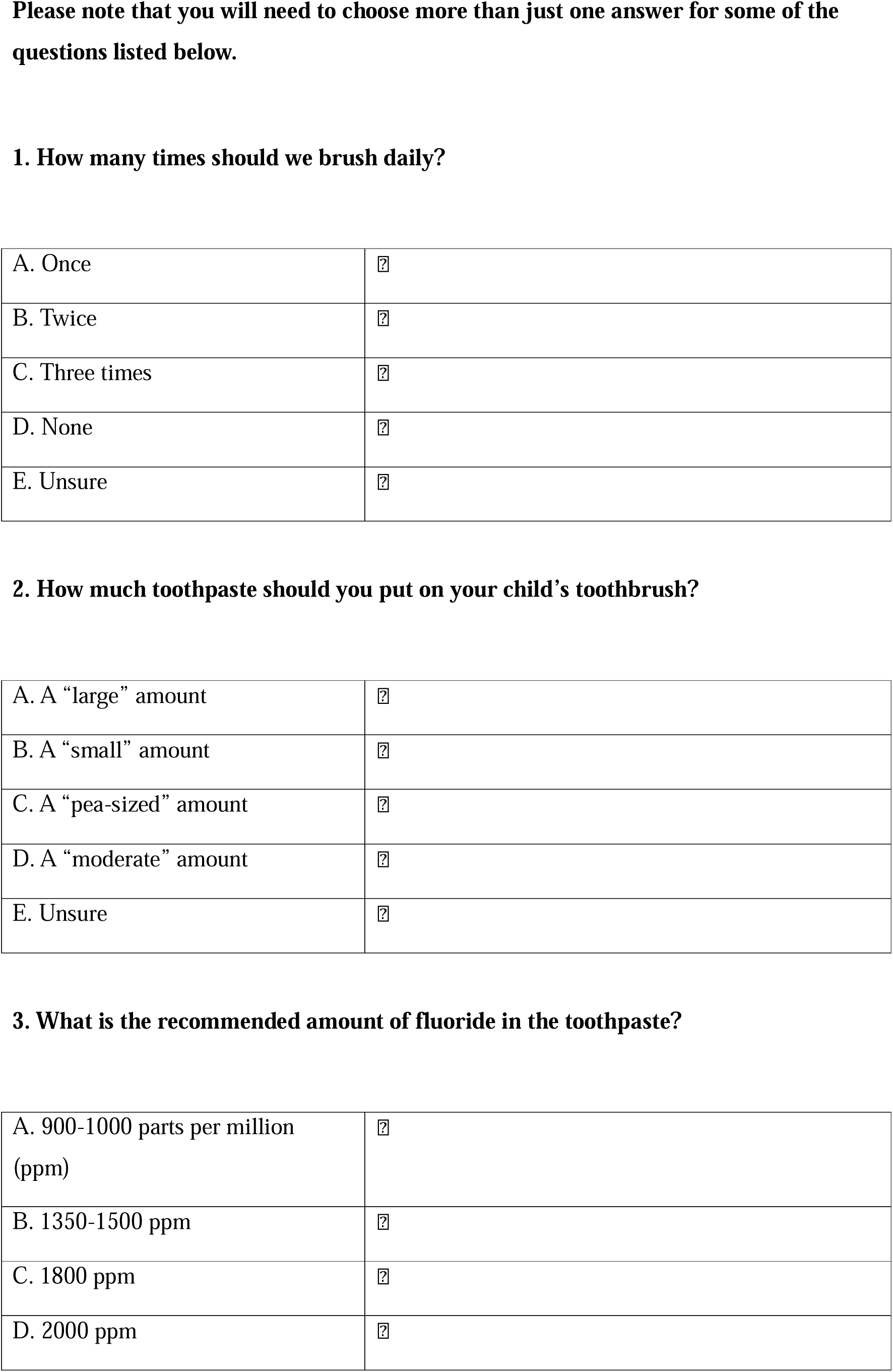

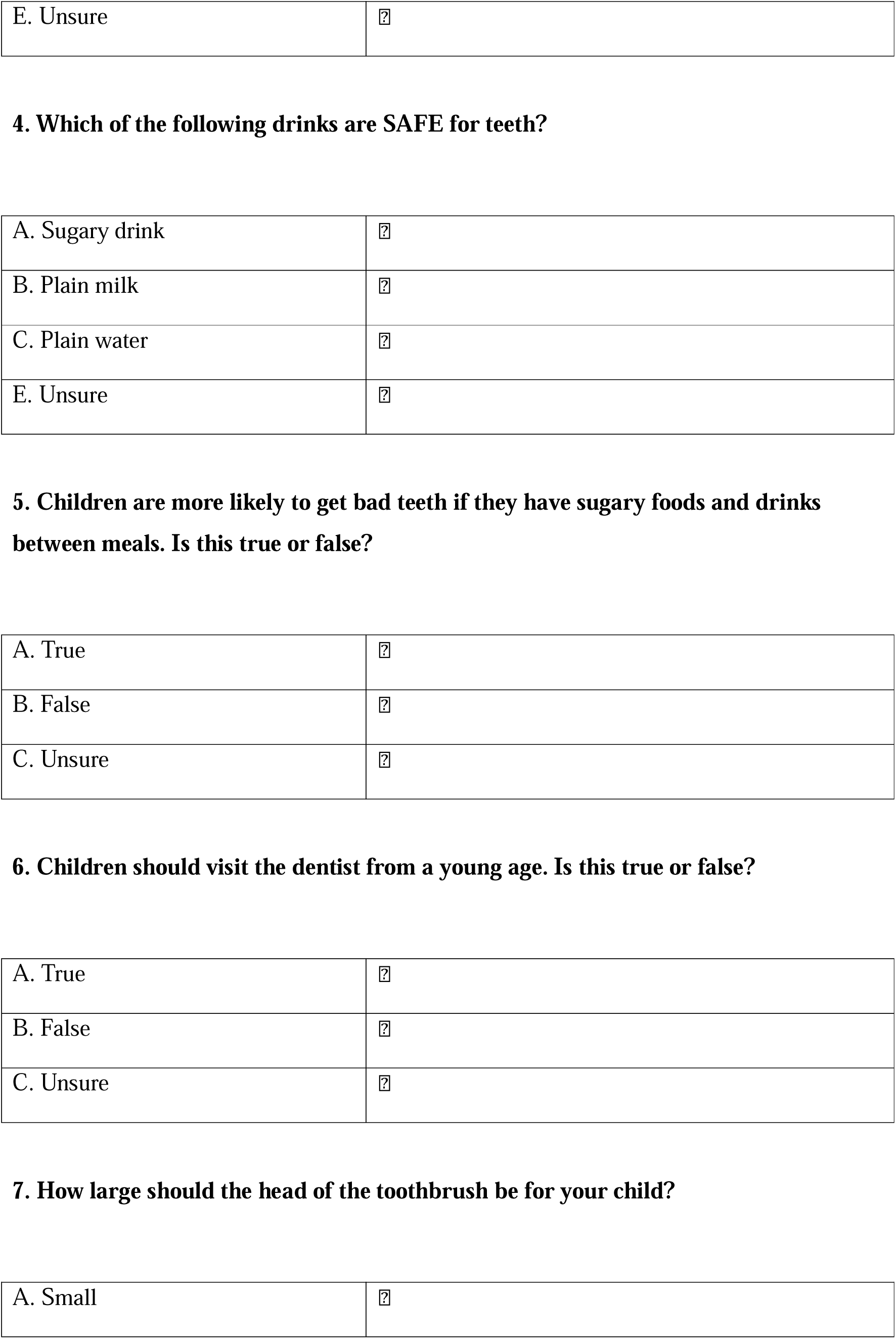

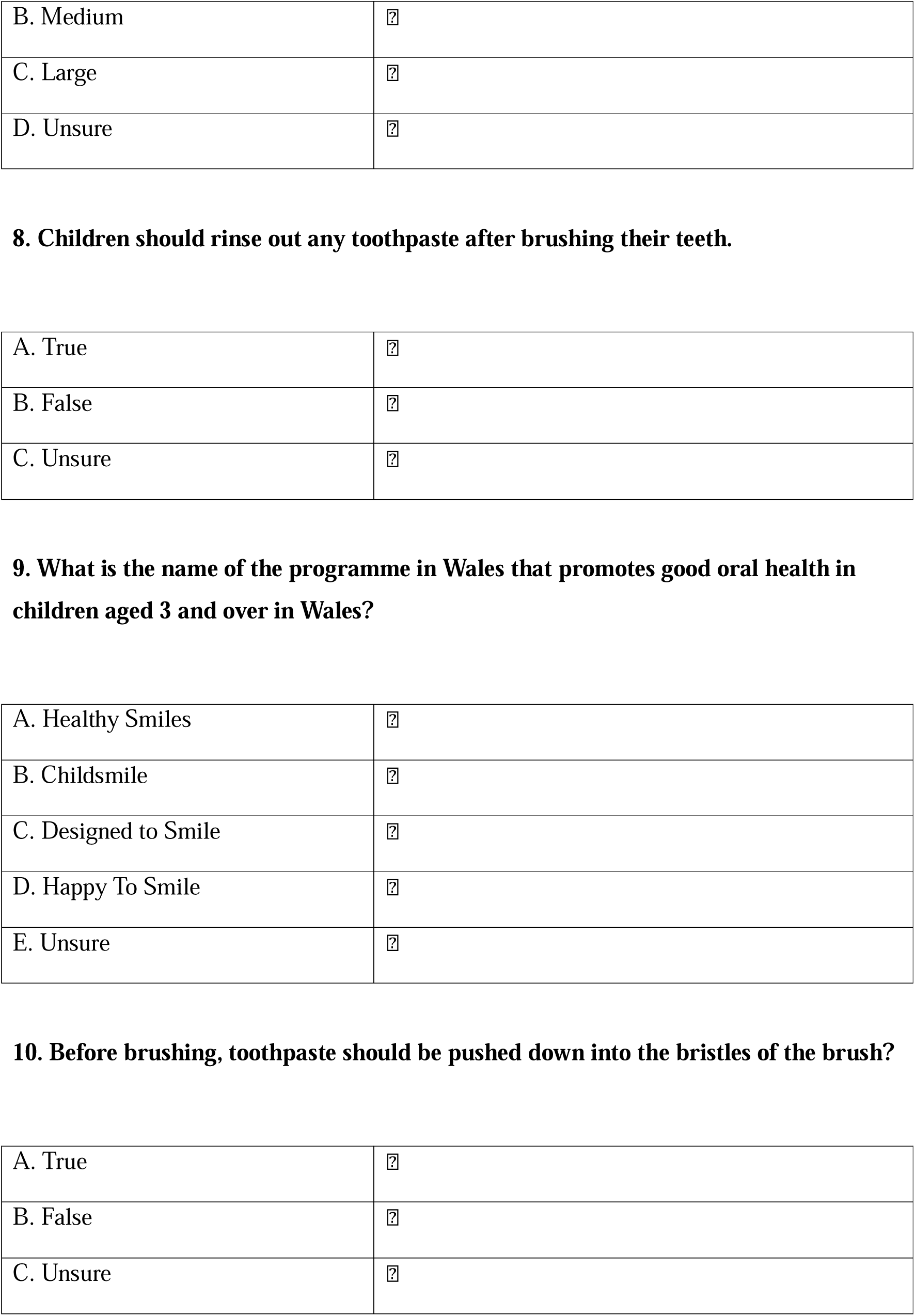

## Notes

### Competing Interest Statement

The authors have declared no competing interest.

### Author Declarations

Ethical approval was granted for this study in April 2022 from the Dental School Research Ethics Committee at Cardiff University, UK (project number: 2204).

### Funding

- Institute of Physics and Engineering in Medicine (IPEM)

